# The experiences and impact of the COVID-19 pandemic on Young Carers: practice implications and planning for future health emergencies

**DOI:** 10.1101/2023.10.25.23297548

**Authors:** D Hayes, D Fancourt, A Burton

## Abstract

**Background:** Young Carers faced significant challenges brought on by the COVID-19 pandemic. We explored the impact of the pandemic and associated restrictions on mental health, wellbeing and access to support in Young Carers in the United Kingdom (UK) to understand how to improve services, as well as support this population in future health emergencies.

**Method:** We conducted 22 qualitative semi-structured interviews from May to November 2021, with 14 Young Carers and 8 staff working in organisations that supported them. Interviews took place remotely over video or telephone call, discussing topics such as experiences of the pandemic on their health, wellbeing and caring responsibilities. We used reflexive thematic analysis to analyse interview transcripts.

**Results:** We identified 4 overarching themes pertaining to the impact of the pandemic and associated restrictions on mental health, wellbeing, and access to support in Young Carers in the UK: 1) challenges to following the guidelines, 2) changes to and loss of routine, 3) changes in provision of informal and formal support and 4) better understanding of inner resilience and goals. Many participants struggled with their mental health and wellbeing as a result of pandemic related restrictions, impacting on support structures for themselves, as well as the individual cared for. However, positive impacts pertained to additional support from local authority and third sector organisations.

**Conclusions:** Our findings highlight some of the changes that affected Young Carers during the COVID-19 pandemic. The impact of changes to routine and a reduction in pre-pandemic support were the greatest concerns reported by participants in this study. The additional support provided by local authority and third sector organisations during social restrictions suggests such organisations could play a greater role in supporting this population going forward, and that schools and Governments may wish to put in additional strategies and provisions to protect this population in the future.

## Background

Prevalence studies highlight that 7-8% of children and young people across Europe undertake caring responsibilities (Carers Trust, 2016; Istat, 2023; Joseph et al., 2020; Leu et al., 2019; van Tienen et al., 2020). In the United Kingdom (UK), there are more than one million young people who are classified as ‘Young Carers’ (Carers Trust, 2016, 2020). These individuals provide substantial care- giving assistance to a family member or friend with a physical or mental illness, disability or a substance misuse problem (Cheesbrough et al., 2017; Leu & Becker, 2019). Referred to as a ‘hidden army’ (Leu et al., 2022; Martin, 2021; Stamatopoulos, 2015), Young Carers take on multiple and varied responsibilities, including delivering personal care, support with activities of daily childcare, and assistance with healthcare appointments (Chadi & Stamatopoulos, 2017; Stamatopoulos, 2015, 2018). However, the work of Young Carers is often invisible to statutory organisations (James, 2017), meaning their needs are not met on a day-to-day basis, and even less so, during national and international health emergencies.

### The role of Young Carers

Estimates suggest that the majority of Young Carers are in their early teenage years (Children’s Commissioner, 2016). This age is a time of important biological, psychological, and social change for the individual as they enter adolescence and become a young adult (Becker & Sempik, 2019; Levine et al., 2005). For most young people, the period of adolescence is marked by forming closer social connections to their peers, developing their own interests and sense of identity, and starting on a path of navigating their own independence (Steinberg & Morris, 2001). However, this is not the case for many Young Carers, who by virtue of their role, are unable to share and engage in these experiences to the same degree as their non-carer peers (Leu & Becker, 2019). This too, can extend into their early adult years, as they may need to take their caring responsibilities into account when making choices around higher education or employment (Leu & Becker, 2019).

Caring responsibilities also have a significant impact on mental health and wellbeing; around half of Young Carers report stress and mental health difficulties associated with caring (Becker & Sempik, 2019). In a meta-analysis, children and young people with chronically ill parents in need of caregiving, were more likely to have high rates of both internal and external behavioural problems, with larger effects when the family unit were of a lower socio-economic status, the illness was longer in duration, and when the child or young person was younger in age (Sieh et al., 2010). Moreover, externalising difficulties were greater when a higher proportion of mothers were unwell and when the young carer was supporting a single parent (Sieh et al., 2010). These difficulties may persist into adulthood, with Young Carers more likely to be hospitalised due to their mental health difficulties and to report feelings of suicidal ideation (Hovstadius et al., 2015).

### Young Carers during the COVID-19 pandemic

The Covid-19 pandemic disproportionally affected young people’s mental health compared with other age ranges (Fancourt et al., 2021). Specifically for children and young people, higher rates of depressive symptoms and lower rates of life satisfaction were found among children and young people during the pandemic compared to a matched control the year before (Mansfield et al., 2022). Reasons for these changes included educational disruption and school closure affecting routine and the ability to interact and socialise with peers (Lee, 2020; McKinlay et al., 2022), increased uncertainty about the future at a time of key developmental milestones (Ganson et al., 2021) and an increase in external stressors, including bereavement (Young Minds, 2021). Research exploring the impact of the pandemic on Young Carers suggests poorer mental health and wellbeing outcomes compared to those who do not have caring responsibilities. For example, a longitudinal survey in Italy observed higher rates of mental health difficulties, including anxiety, depression and wellbeing, as well as higher levels of Covid-19 specific outcomes; risky health behaviours, loneliness, home violence, and fear of Covid-19 in young people with caring responsibilities compared to those without (Landi et al., 2022). Similarly, data from the Millennium Cohort study in the UK showed that Young Carers had higher psychological distress and lower mental wellbeing than their non-carer counterparts during the early stages of the pandemic (Nakanishi et al., 2022). Psychosocial risk factors including poor quality sleep, low social support, and increased feelings of loneliness, largely explained findings.

Qualitative findings also lend support to the role of psychosocial risk factors during Covid-19, as well as shedding further light on the impact the pandemic had on children and young people with caring responsibilities. For example, a rapid approach, mixed-methods study identified that during the pandemic, support for Young Carers was withdrawn from external providers as well as friends and family, often leaving Young Carers overwhelmed, unable to cope, and without coping mechanisms and routines (Blake-Holmes, 2020; Blake-Holmes & McGowan, 2022). The loss of school as a sanctuary and source of support was highlighted, as well as not being able to have time and space to manage their mental health difficulties. Other studies have identified similar themes (Martin, 2021; Wylie-Curia, 2021), as well as reporting of positive experiences associated with the Covid-19 pandemic, including more time for self-care and more time with family (Martin, 2021; Wylie-Curia, 2021).

### This study

Young Carers are referred to as a ‘hidden army’ and are often invisible to statutory organisations. This not only means that their physical and psychological needs are frequently not met in day-to-day life, but that in times of emergency, the exacerbation of these needs can be overlooked. Qualitative research on this population and the Covid-19 pandemic has mostly focused on the initial and early stages of the pandemic (Blake-Holmes, 2020; Blake-Holmes & McGowan, 2022; Martin, 2021; Wylie-Curia, 2021), is often limited to very small sample sizes (Martin, 2021; Wylie-Curia, 2021), and only focuses on the views of the Young Carers (Wylie-Curia, 2021) rather than sourcing multiple stakeholder views, such as those running services. This study, therefore, explored the experience of Young Carers during a global health emergency, with the aim of developing strategies for the protection and support for Young Carers during future health emergencies.

## Methods

### Design

A qualitative interview study was undertaken to explore the impact of the Covid-19 pandemic and associated restrictions on mental health, wellbeing and access to support in Young Carers aged 13 to 24. The work forms part of the [REMOVED DUE TO BLINDING] study.

### Participants

Participants were recruited via national and local third sector organisations providing services and support to Young Carers, social media and via a study newsletter. Both Young Carers aged between 13-24 years old and staff working for organisations that supported Young Carers were eligible to take part in the study. Participants also had to be based in the UK, be able to provide informed consent, or assent alongside parent/guardian informed consent if aged 13-15 years old. Study posters containing researcher contact details were sent to UK based organisations providing services to Young Carers for distribution to their staff and Young Carers via newsletter/mailing lists. A researcher (INITIALS REMOVED FOR BLINDING) also attended online and in-person (when restrictions allowed) young carer support groups provided by these organisations to introduce the study and answer questions about the research. For those that participated in an interview, a £10 voucher was provided for their time.

### Data collection

Interviews took place between May and November 2021. Semi-structured topic guides were used to conduct the interviews. For Young Carers, questions explored the impact of the Covid-19 pandemic and associated restrictions on mental health, wellbeing and access to support. For staff, questions included the impact of the Covid-19 pandemic and associated restrictions on service user mental health and wellbeing, challenges to operational practices and service delivery and service adaptations. [see Supplementary Information A for the topic guides]. Interviews were conducted remotely either via telephone or Microsoft Teams. Interviews with Young Carers ranged in length from 37 to 69 minutes (*M = 52, SD 11)* and for service providers between 36 to 66 minutes (*M = 48, SD 13)*. All interviews were conducted by [INITIALS REMOVED FOR BLINDING], a [GENDER REMOVED FOR BLINDING], Senior Research Fellow with substantial experience in qualitative research methods and mental health research. All procedures involving human participants were approved by the UCL Ethics Committee (project IDs: 14895/005 and 6357/002).

### Data analysis

Audio files were transcribed verbatim by an external transcription company approved by UCL. Personal and identifiable data were removed from transcripts before analysis to maintain confidentiality. Transcripts were then imported into NVivo 12 (QSR International Pty Ltd., 2018) for data management and subsequent coding. A reflexive thematic analysis was conducted (Braun et al., 2022). First, initial codes were developed based on the topics explored in the interview schedule.

Codes were then applied to relevant fragments of text within the transcripts, however new codes were also developed when passages of participant accounts did not align with the pre-determined coding framework. Coding was undertaken primarily by [INITIALS REMOVED FOR BLINDING] with additional coding support provided by [INITIALS REMOVED FOR BLINDING]. The coding framework was discussed and refined during the data analysis period within regular meetings between the research team. Codes were then organised into groups of similar constructs and labelled with overarching theme and subtheme names, with [INITIALS REMOVED FOR BLINDING presenting findings to the research team for discussion and refinement at two follow up meetings. Revisions to findings were made based on feedback at these meetings and a final set of themes and sub themes were agreed.

## Results

14 Young Carers and 8 staff working for Young Carer organisations participated in an interview for this study. The mean age of young carer participants was 19 years old and the majority were female (71%) and lived with family (93%). The largest proportion of Young Carers identified as ‘Asian Bangladeshi’ (43%) and were at school or university (36%). Parents were the most frequent recipient of care from the young person (43%).

The mean age of representatives from service providers was 39 years old with, on average, 7 years of experience working with Young Carers. All identified as White and outlined that the services they provided included advice and information, as well as signposting to services.

Demographics of participants are presented in Tables 1 and 2.

**Table 1:** Young carer characteristics.

| **Characteristic** | **Mean (range)/ n(%)** |
| --- | --- |
| **Age** | 19 (14-24 years old) |
| **Gender** |  |
| Female | 10 (71%) |
| **Ethnicity** |  |
| Asian Bangladeshi | 6 (43%) |
| White British | 3 (21%) |
| Asian Pakistani | 2 (14%) |
| Mixed Race – White and Black/Black British – Caribbean | 1 (7%) |
| Black/Black British – African | 1 (7%) |
| White other | 1 (7%) |
| **Employment status** |  |
| At school/university | 5 (36%) |
| At school/university and part time employment | 4 (29%) |
| Part time employed | 3 (21%) |
| Full time employed | 1 (7%) |
| Unemployed and seeking work | 1 (7%) |
| **Living situation** |  |
| With family | 13 (93%) |
| Live alone | 1 (7%) |
| **Who they provide care to** |  |
| Parent | 6 (43%) |
| Grandparent | 3 (23%) |
| Multiple family members | 3 (23%) |
| Sibling | 1 (7%) |
| Friend | 1 (7%) |

**Table 2:**
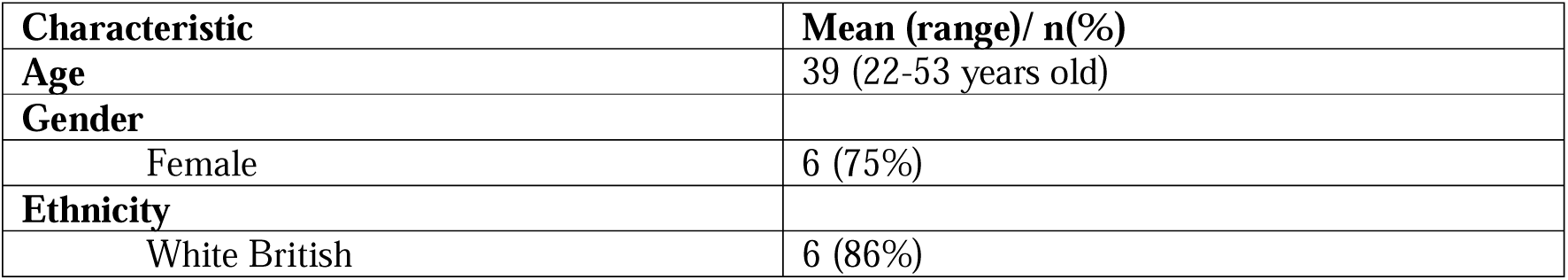
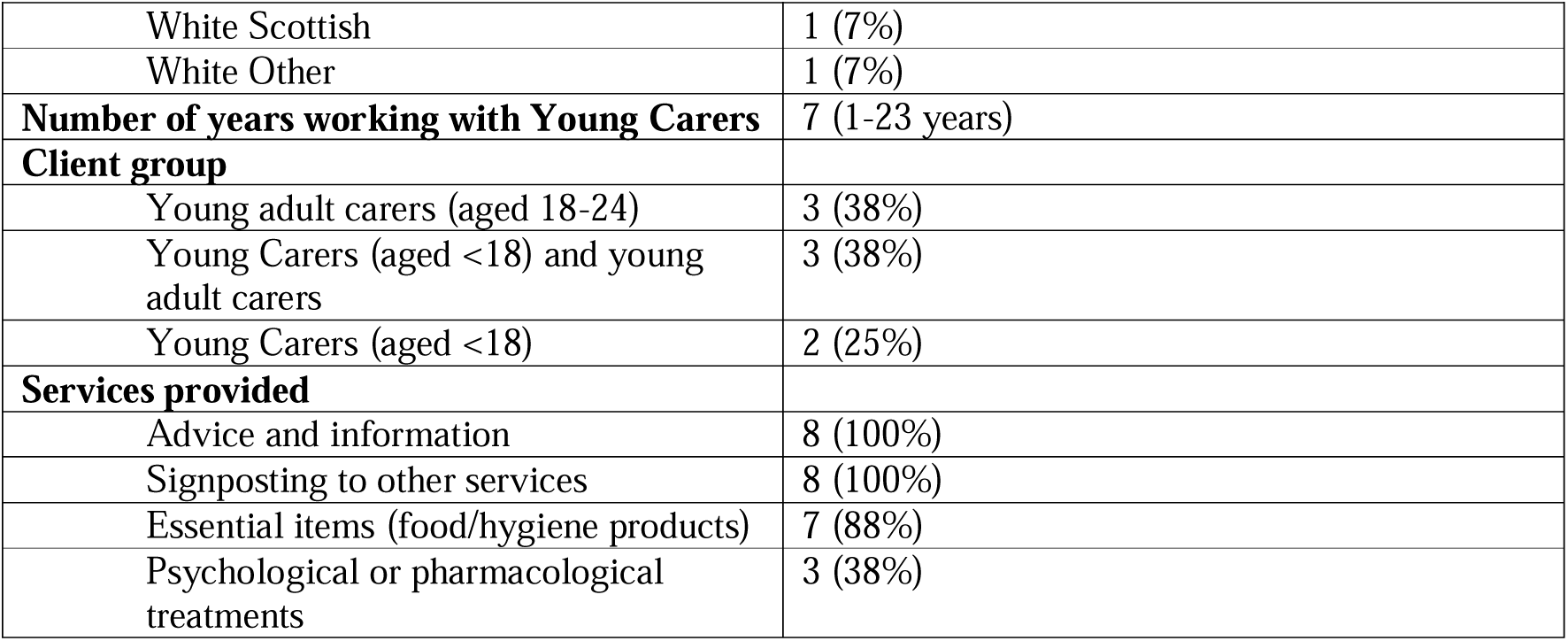
Service provider characteristics.

### Themes

We identified 4 overarching themes pertaining to the impact of the pandemic and associated restrictions on mental health, wellbeing, and access to support for Young Carers in the UK: 1) challenges to following the guidelines, 2) changes to routine, 3) changes in provision of informal and formal support and 4) better understanding of inner resilience and goals. Themes and sub themes are illustrated in Figure 1 and described in detail below with supporting quotations from participants.

### Theme 1: Challenges to following the guidelines

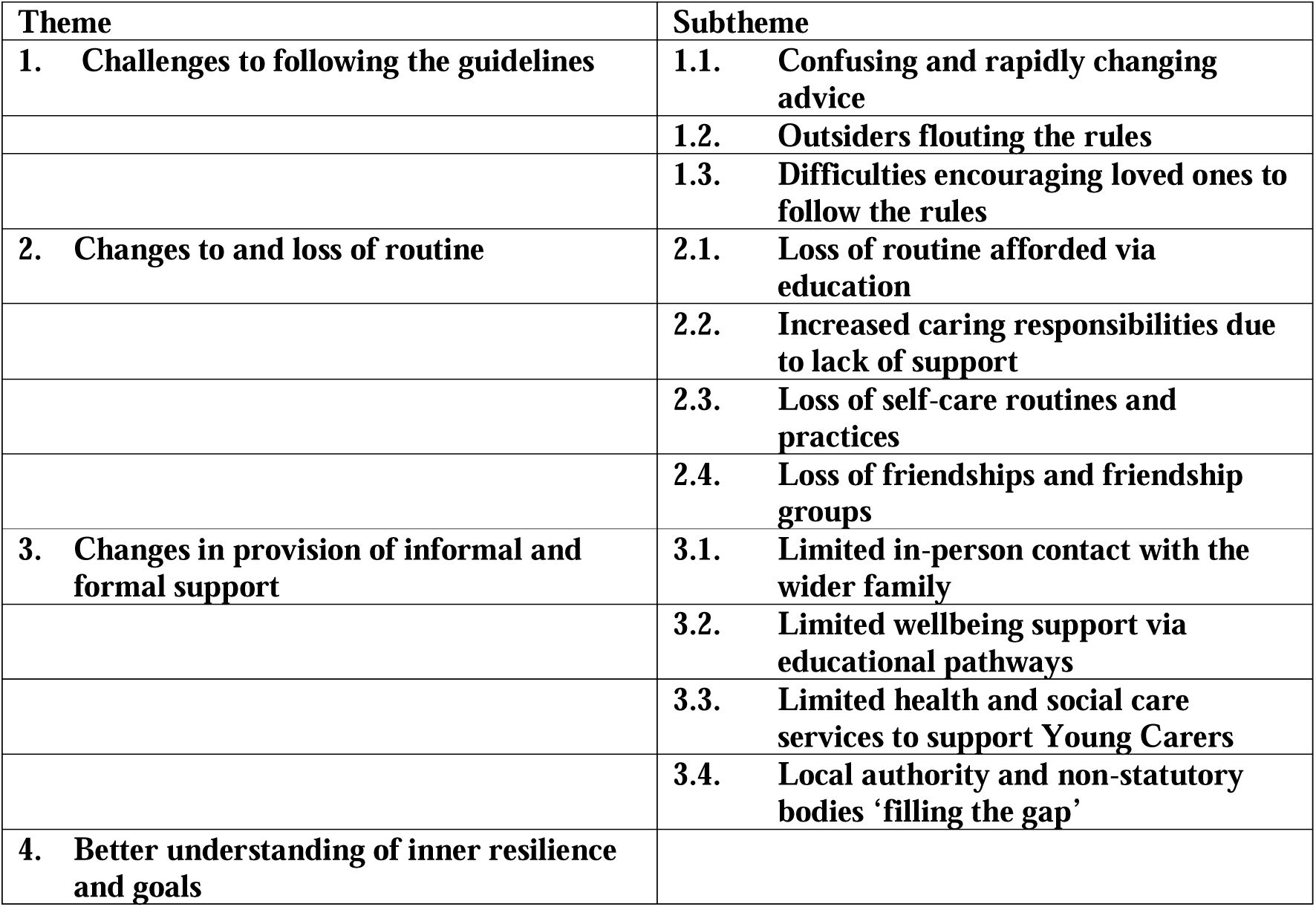

Young Carers outlined various challenges when it came to both them and others being able to understand and follow the UK national government rules and social distancing guidelines:

> *‘[It] was quite difficult because not many people really knew what was going on of course. We had periods where we didn’t have to wear masks, periods where we wore masks all day every day’ (Young Carer 6, age between 14- 17, looks after grandparent)*

#### 1.1 Confusing and rapidly changing advice

The majority of Young Carers described the importance of following the guidelines, understanding that they were put in place to protect themselves and the public by limiting exposure and transmission of the virus. In line with this, many Young Carers attempted to follow Government advice and take precautions via methods such as: self-isolating, forming support bubbles, getting vaccinated, social distancing and wearing masks.

> *‘I did self-isolate, and it was to protect both myself and my mum from the virus, and I didn’t want to be a carrier and then spread it’ (Young Carer 9, age between 18-21, carer for parent)*

For some, taking these precautions was intrinsically linked to their role as a Young Carer, where they felt a strong sense of duty to protect the person they were looking after due to them having an increased vulnerability to the virus.

> *“I feel as though if I wasn’t a carer, I probably wouldn’t think of the guidelines as something quite serious, but because I am, I have to think about not only myself, but the people I’m caring for. So, that motivates me to follow the guidelines properly, wear a mask, be socially distanced” (Young Carer 1, age between 18- 21, carer for multiple family members)*

Despite the general consensus that following the guidelines was a good idea, some Young Carers reported that they lacked clarity or sense, and were therefore difficult to follow:

> *“In terms of the tiers, it was a bit all over the place, because one minute we were in Tier 2, then we were Tier 3, then we had a new tier, we were in Tier 4. So, it was quite unnerving, because we weren’t sure where we stood” (Young Carer 7, age between 18- 21, carer for parent)*

This left Young Carers feeling confused, frustrated or anxious, as at times, there was discordance between official guidelines and how they felt they, and others, were supposed to act to help stop the spread of the virus and protect themselves and others.

> *“Even though social distancing and everything has eased it’s like further than that okay I need to maintain my distance, just so we’re not spreading anything to one another constantly” (Young Carer 3, age between 18- 21, carer for parent)*

#### 1.2. Outsiders flouting the rules

Some Young Carers reported tensions with others, who they perceived as not being diligent or stringent in following the guidance. In most cases this pertained to members of the general public, but also to acquaintances and friends, particularly at school. Young Carers felt let down by others and perceived their actions as selfish and putting vulnerable people at risk:

> *“I [felt] quite a lot of annoyance, particularly at people. They just haven’t properly thought about what they’re doing, they went and did something, and they haven’t thought about how it can impact people” (Young Carer 14, age between 14- 17, carer for parents)*

In the latter stages of the pandemic, as Young Carers returned to in person lessons, school was often a place of tension for Young Carers, as they perceived their peers as lax with following the guidelines, particularly mask wearing:

> *“There are some people [at school] who don’t always wear their masks, as in pull them down a bit. I don’t with mine, except to drink” (Young Carer 12, age between 14- 17, looks after brother and mother)*

Sticking to the rules when others were not, sometimes resulted in Young Carers feeling depressed, resentful, angry, and lonely:

> *“But then, when I’ve seen people outside [the house], I’ve been like, oh, they look like they’re having a fun time, and there is a risk that they could catch Coronavirus, at least they are taking the approach that if I go, I go happy, rather than being inside, miserable.” (Young Carer 7, age between 18- 21, looks after parent)*

Service providers also reported hearing concerns from Young Carers around peer non-conformity to the guidelines when attending school and the anxiety that resulted from this:

> *“[They] were anxious at going to school where people weren’t socially distancing and following the guidelines, so they felt vulnerable that they were maybe taking the covid back home to their vulnerable families” (Service Provider 7)*

#### 1.3 Difficulties encouraging loved ones to follow the rules

Whilst it was more often those outside the immediate family unit who were less likely to be following the rules and guidelines, some Young Carers also discussed how their family members also found abiding by the guidelines a challenge. When this involved the person the Young Carer supported, the individual being looked after was often unable to follow the guidelines due to a lack of cognitive capacity, or because of mental health difficulties:

> *“I think she does need some guidance and support, especially when it comes to stuff like wearing a mask, sometimes, she can be quite on and off with that, and she doesn’t like wearing one, especially with her OCD. She actually does not like it when it touches her face” (Young Carer 9, age between 18- 21, looks after parent)*

Conversely, when it was other individuals in the family unit not following the rules, this was often because they didn’t believe in Covid 19, or that they believed they were at a low risk of catching the virus. In such instances, Young Carers reported talking to their family members about this, which often resulted in heated arguments:

> *“[It causes]very, very heated arguments…I begin talking to her about, okay, you shouldn’t be going to your auntie’s house, you should be keeping to yourself right now, you need to be at home, you have to wear your mask and things like that. And it starts going, you know what, I regret having you, and that kind of stuff.” (Young Carer 13, age between 18- 21, looks after parent)*

In such instances, participants reported feeling avoidant with family members when it came to discussing this topic:

> *“I think I’ve tried to have discussions about it, but it just turns out to be an argument, so I don’t really bother very much anymore…I try to avoid the subject” (Young Carer 11, age between 14- 17, looks after parent)*

### Theme 2: Changes to and loss of routine

The pandemic brought changes to the day-to-day routines of individuals in the UK. For Young Carers, this meant a loss to some important aspects of their day, such as time with friends at school, which provided a break from caring. Importantly, routine, structure, and having things to do were acknowledged as being important by Young Carers and Service Providers in supporting Young Carers wellbeing. Whilst structures and routines varied depending on the individual, rules brought in to combat the pandemic meant that much, or all, of the routines Young Carers utilised to support their mental health disappeared, leaving them stuck in limbo:

> *“By getting up in the morning, you go out, you might tidy your house, you might interact with friends, colleagues, you occupy yourself in some way, and those daily experiences keep you stimulated and engaged with life in some way. They don’t have that. They’re sort of morbidly stuck, which leads to a psychological stuck-ness” (Service Provider 8)*

#### 2.1 Loss of day-to-day routine afforded via education

For many Young Carers, the routine provided by school was a welcome part of the day, allowing them to focus on themselves, their education and provide some respite from caring responsibilities. However, the scramble for teaching staff to adapt lessons to online formats due to the pandemic, often left ‘gaps’ in the school day, removing the structures that many Young Carers relied on and leaving an emptiness:

> *“So when lockdown happened and I had nothing to revise for, no studying, no pre-reading, no nothing. I’m just left with this endless void of time” (Young Carer 13, age between 14- 17, looks after parent)*

The knock on consequences were that many worried about how this would impact their education and future prospects:

> I’m worried about if I’m going to have enough time to focus on my GCSEs compared to how much I need to care for my nan. Because it’s going to get more intense as years go on or months go on even. And trying to do that and GCSEs is a lot (Young Carer 6, age between 14- 17, looks after grandparent)

#### 2.2 Increased caring responsibilities

The majority of Young Carers described how the pandemic increased their caring responsibilities and the burden associated with caring. The dual set of circumstances of not having access to previously available sources of support (see Theme 3) and not being able to go to school or work meant that Young Carers felt obligated in taking on additional responsibilities to secure the needs of the person they cared for. Many Young Carers reported this as challenging:

> *“In terms of my responsibilities, I would just say they just became a bit more demanding, because, obviously, I couldn’t leave the house, and things like that. So, it was quite full-on, because we were at home 24-7, so just had a lot more responsibilities in the house…I had to cook more often, I had to clean more often. Where, before, we could go to the community centre and do community activities and stuff like that” (Young Carer 4, age between 22- 24, looks after parent)*

For some, this increase in caring responsibilities meant clashes with other priorities such as their education. Instead of having protected time away from home to study, being stuck at home and therefore constantly available, resulted in taking on additional tasks and disrupted learning which caused individuals to fall behind:

> *“I think it was just difficult because I couldn’t balance school and having to look after my mum at the same time. I’m not sure how to word it properly. It was just difficult because I felt like I couldn’t really focus in school because I had to look after my mum because I was at home. So I just had to focus more on my mum rather than schoolwork” (Young Carer 11, age age between 14- 17, looks after parent)*

For others, being stuck at home and increased caring responsibilities meant that, for the first time, they became aware of the nature and level of the difficulties of the person they cared for, which resulted in them feeling scared;

> *“Yes, it was, because when I’m at uni, obviously, I’m not seeing him all the time. But at home, 24-7, I’m seeing things that I didn’t see before…His feeding, for example, I’ve noticed that he actually chokes a lot more than usual. Partly when he has difficulties breathing, and that scares us, because he has an DNR, so it’s do not resuscitate, but, we get scared, if something were to happen, we can’t do anything about it. I never used to see that before, because I was always at uni during his feeding, but now that I’m home, I’m seeing it often, and he actually scares us a lot” (Young Carer 1, age between 18- 21, look after multiple family members)*

However, for other participants, whilst caring responsibilities increased, not having to go to school or University, or having a more adaptable education or work schedule brought a welcome degree of flexibility to their routine. This was often the case when they were not the sole person in charge of care, or when the individual they cared for had greater independence, which left more time to carry out additional responsibilities and resulted in fewer stressors:

> *“It’s been quite good [pandemic changes], actually, [Name], because, obviously, I’m still able to do my caring responsibilities. I take regular breaks and I take my lunch break every day. I also take regular breaks throughout the day, we’re encouraged to take breaks throughout the day, so I just do some cooking, some cleaning, and anything else, whether mum’s got a doctors appointment she needs to book, or things like that, really, I’m able to do them” (Young Carer 4, age between 22- 24, looks after parent)*

#### 2.3 Loss of self-care routines and practices

Each individual spoke about needing their own time and space before the pandemic as a form of self-care, to help manage the demands of being a Young Carer; these varied in nature, but included hobbies, sports and creative outlets and often involved being outside the home. As restrictions were bought in, this limited Young Carers ability to engage in certain activities that supported their mental health such as sports or going to the gym:

> *“When I stressed out… I’ll go to the gym, go for a walk, have a workout, but during lockdown, everything was closed, so I didn’t get that chance” (Young Carer 1, age between 18- 21, looks after multiple family members)*

Not being able to leave the house, due to pandemic restrictions, as well as increased caring responsibilities, meant that Young Carers were unable to find time to cope and decompress. This led to feelings of isolation, low mood, as well as feeling trapped and overwhelmed:

> *“Sometimes when I’d have mood swings they would be triggered by anyone just coming in my personal space when I’m trying to have that alone time, and some of it would just be so random just because I’m not able to go out as well” (Young Carer 3, age between 22- 24, looks after parent)*

#### 2.4 Loss of friendships and friendship groups

In addition to relying on hobbies and other activities, Young Carers also spoke about the importance of friends as a source of coping with the demands of caring. Having friends and being part of friendship groups meant they could enjoy similar experiences to their peers, such as hanging out and going to the movies. The pandemic restrictions meant that most of these activities were limited to digital or video format. This resulted in friends spending less time together as the pandemic took its toll on people’s routines and health:

> *“Friends that I already knew, we saw each other much less, because everyone had a lot of things on their plate. Either, I had a friend who had a lot of things going on with work and their own studies or other friends had things going on with their mental health and the pandemic, and then I had things going on with work, mum’s health, the housing situation, and my studies. So, everyone having a lot of things going on, it was just barely, really, seeing each other “(Young Carer 2, age between 22- 24, looks after parent)*

For many this decrease in contact led to friendship groups fragmenting and a loss of a source of coping for Young Carers:

> *“I was in touch with a few more [friends] before the pandemic. I’m still in contact with a few now, but it’s sort of dribbled out and stopped now because I think it’s difficult trying to arrange to meet up” (Young Carer 14, age between 14- 17, looks after multiple family members)*

For some Young Carers however, these feelings were tempered by an acknowledgment and realisation that either they had changed and were not the same person as prior to the pandemic, or that the people they classed as friends were only due to proximity, rather than shared values or hobbies:

> *“So, we all noticed that mostly we had nothing in common, over the lockdown we’d all changed so much we just had nothing in common anymore” (Young Carer 6, age between 14- 17, looks after grandparent)*

Following this realisation, Young Carers stressed the importance of being around people that ‘mattered’ or who were more aligned to their own interests:

> *“I don’t think so. We don’t really talk as much anymore, so we just stopped being friends….it did kind of make me realise that I don’t really want to stick with this person” (Young Carer 11, age between 14- 17, looks after parent)*

### Theme 3: Changes in provision of informal and formal support

To help manage caring responsibilities before the pandemic, many Young Carers told us that they relied on both formal sources of support, such as healthcare services and Young Carer organisations, as well as informal sources of support, such as family members. Rules around how individuals were allowed to interact meant such provisions drastically changed, with limited contact from wider family members, reduced health and social care support, and limited emotional support provided by schools. However, some third sector agencies provided extra or new support, via financial incentives and food packages.

#### 3.1 Limited in-person contact with the wider family

Prior to the pandemic, some Young Carers were supported in their roles by other members of their extended family:

> *“I’ve got relatives that do live locally… so during my second year, prior to the pandemic, these relatives were very supportive with me, because at that time, I couldn’t really manage my caring responsibilities” (Young Carer 9, age between 18- 21, looks after parent)*

As rules and guidelines limited contact between different households, this meant that many Young Carers lost this additional support and had to take on extra responsibilities:

> *“When I used to go to Uni, I had my uncle or someone at home, so they would do some of the things, like helping feeding at lunchtime. Because, now, I’m home 24-7, I don’t get that extra help, because they’re in their house during lockdown. So, I’m having to do more things throughout the whole day, rather than just bits and bobs here and there” (Young Carer 1, age between 18- 21, looks after multiple family members)*

Not having this support left Young Carers feeling overwhelmed and stressed as they were often the only individual managing their family member’s care and felt they were always on call:

> *“It was mentally and physically draining, because you’re not just having to think about doing this for an hour, it’s 24-7” (Young Carer 1, age between 18- 21, looks after multiple family members)*

#### 3.2 (Limited) wellbeing support via educational pathways

Educational settings not only provided routine and structure, but also acted as a conduit for accessing mental health support from staff, as well as more formal support from child and adolescent mental health services. Some Young Carers reported that there was less support available than previously, due to the system being overloaded, with staff juggling changes to learning and an increased need for support as the pandemic worsened mental health among young people more generally:

> *“it’s someone to just chat to if you needed them. Also not just outside of school, so inside of school there were counsellors on hand ready if you needed them, and it was just helpful sometimes to have someone to go and talk to. But lots of that has been put on pause during the COVID situation” (Young Carer 14, age between 14- 17, looks after multiple family members)*

When these support structures were not in place, Young Carers reported feeling anxious, low, depressed, struggling to cope, and let down. However, for Young Adult Carers in further education, or for those whose Young Carer status was known by teachers in statutory education, support was more readily available:

> *“I had [support], actually because of Young Carers I had a teacher that was set to me because I was a young carer. They would email me once a week just to ask how I was” (Young Carer 6, age between 14- 17, looks after grandparent)*

Whilst this support was welcomed, for those that received it, there was a feeling that it was limited in scope, or that it came too late:

> *[It took] two or three months, to actually send an email and give me some kind of assessment. But by that time, it was too late, because then the lady was like, you’re not going to be a student anymore, because I will be graduating soon, so I can’t really be eligible for the service anymore” (Young Carer 9, age between 18- 21, looks after parent)*

#### 3.3 Limited health and social care services to support Young Carer

Prior to the pandemic, many Young Carers relied on the support of different health and social care workers to help facilitate the care of the person they looked after. This included care workers who would help with personal hygiene and daily living, as well as nurses and other healthcare professionals.

As restrictions and social distancing measures were put in place, this meant that some home visits were stopped, scaled back, or used only to provide essential care, rather than additional support:

> *“So, yes, the support had been minimised a bit. We have carers coming in and out, throughout the day, to change him, but other than that, if you were to request extra support, it would go on hold, because there are other priorities, or it’s because of lockdown, they’re trying to minimise them coming into our house or us going to the hospital” (Young Carer 1, age between 18- 21, looks after multiple family members)*

It also meant that in-person health and social care appointments, both for the Young Person and the person they looked after, were cancelled or rescheduled unless perceived as an emergency:

> *“Yes, appointments have been less frequent. Non-essential ones like the dentist, unless you had an urgent problem, that was put on pause, and when people did come round they’d be fully clad in PPE and looking a bit like some otherworldly creature. But yes, not a great deal, because it wasn’t too frequent anyway” (Young Carer 5, age between 22- 24, looks after parent)*

Whilst there was an acknowledgement by Young Carers that health and social care staff were doing the best they could, the use of remote appointments were experienced as inappropriate in some instances, particularly for assessments of physical health complaints:

> *“The issue of needing to see someone, so they could assess her feet and her legs, you can’t really do that over the phone” (Young Carer 7, age between 18- 21, looks after parent)*

> *“Obviously, not being able to see my mum physically, the doctors, they couldn’t really see her physical state, especially with her weight…” (Young Carer 9, age between 18- 21, looks after parent)*

This caused worry and was experienced as substandard care with health needs not properly addressed. Moreover, in many instances, Young Carers felt dismayed and hopeless as not getting the necessary treatment support resulted in their family member deteriorating and them being left to cope with this on their own:

> *“Having to have mum have treatment was also delayed, having people come out, having short staff, so lots of things were delayed, meetings, and because there wasn’t enough staff or people working at certain times. Things took longer than needed with mum’s health…that was really hard, because there was a point where her mental health was very bad, and so, I had to cope with her mental health being bad, in order to get support. And then, once the support started, it took quite a while” (Young Carer 2, age between 22- 24, looks after parent)*

#### 3.4 Local authority and non-statutory bodies ‘filling the gap’

Whilst some services were unavailable, or not as accessible, due to pandemic related restrictions, other services provided by local authorities and voluntary organisations were able to provide additional or adapted support to participants. Often this was possible via special emergency allocations made accessible for vulnerable groups from local government and charities:

> *“They’ve [Young Carers] been getting all the different supplies for activities from us. Things like the [Charity appeal]. Whenever we’ve been getting funding we’ve been able to give what we think they need” (Service Provider 7)*

In the first half of the pandemic, services reported providing financial support, often to meet the practical needs of the Young Carer and family members, for essentials such as food, bedding, and clothing:

> *We had quite a number of financial support requests near the start, as well. And mid-pandemic…there was a point where we were issuing out Tesco vouchers to try and alleviate some of that financial pressure (Service Provider 3)*

Young Carers reflected that having this financial support, often when they, or their family members were furloughed, out of work, and stuck at home leading to increased food and utility costs, was a lifeline:

> *“So, I think I got £400 from the carers centre. That really supported me, [Name], in terms of getting basic food and drink, but also basic clothing items, even, and just some bedding materials. It was just a really nice gesture from the carers centre, because, obviously, they’d provided that grant, and I was just able to spend it back on my mum and just the family, to see if there was anything I could provide. That really helped and I was just so grateful for that” (Young Carer 4, age between 22- 24, looks after parent)*

Local authorities and other third sector organisations also provided food packages to Young Carers and their families.

> *“I do vaguely remember how we used to have, the council used to drop off food to more vulnerable people, so we got the basics from that, but more stuff like perishables and more snacks, we would allocate one person to get them” (Young Carer 8, age between 18- 21, looks after multiple family members)*

On top of the essentials to survive, services were able to provide Young Carers with equipment to be able to participate in online learning for education and socialising:

> *“half of their houses didn’t have wifi in them, so we were having to go and buy dongles for them that they could get wifi access to be able to come and do a Zoom. When the schools were taking the Chromebooks back…we then applied for so many in our own right, and then I was going around giving them to the families that were most at need” (Service Provider 8)*

Third sector organisations also provided emotional and social support for participants during the pandemic. In its initial stages, this was only possible via videocall. This often provided a dual purpose, allowing Young Carers a forum to support one another and knowing that individuals on the call were in the exact same situation as them:

> *“Even though it was online, it just felt nice, actually, to have a support network, and a group of Young Carers, like myself, who we could share experiences, and I didn’t feel as isolated in my own struggles” (Young Carer 9, age between 18- 21, looks after parent)*

However, some Young Carers found it challenging to engage in these online activities, due to Zoom fatigue and being ‘stuck’ in the environment where they performed their caregiving, which meant that it was seen as another task, rather than a social activity:

> *“And it’s like the whole day of technology at school for me, then having another social event it really didn’t feel like a social event to me it felt more like I’m attending another meeting. Yes, so that was a downside. So for the entire year I didn’t even end up interacting with anyone from the Carers Centre” (Young Carer 3, age between 22- 24, looks after parent)*

Low attendance at online support sessions set up by local organisations to support Young Carers was acknowledged by service providers. Like reports from Young Carers, there was a feeling that ‘zoom fatigue’ from individuals being online all day for educational purposes was contributing to this:

> I know that one of the reasons that the young people didn’t want to engage with us so much was because it was just more screen time, it was just more of this (Service Provider 8)

As pandemic restrictions eased, Young Carers spoke about appreciating the opportunities to meet, in socially distanced ways face-to-face, facilitated by Young Carer organisations and feeling more connected to others in a face-to-face format:

> *“I really enjoy the physical sessions that’s now, because it’s just you can see the whole body, you can see the whole emotion, much better, more clearer…And even if you’re just sitting in silence, I think that humans just want to share being around other people, even if it’s in silence, so I just prefer that much more. Now, I’m attending every single week” (Young Carer 8, age between 18- 21, looks after multiple family members)*

These reports from Young Carers were also echoed by the organisations facilitating contact, highlighting the difference that in-person contact made to Young Carers wellbeing, which couldn’t be replicated online:

> *“We met in the morning, I couldn’t believe it, we stayed until 6 o’clock in the afternoon because I got some picnic lunch. Nobody wanted to go home. It was about nine of us, we were just- okay, you see the animals, because they have the picnic benches and things. We were there for eight hours which I didn’t expect, but they just really craved that- there is also something about the spontaneous moments where like you don’t need focus of conversation. Zoom meetings really can’t be longer than an hour or hour and a half, two hours tops. It’s exhausting” (Service Provider 1)*.

#### 4. Better understanding of inner resilience and goals

When reflecting back on the pandemic and what they had learnt, Young Carers spoke about how it had taught them about their own capabilities, resolve, and resilience in the face of adversity. For some this was related to how they managed to adapt to the increased demands of caring during the pandemic, often with less support than they previously had:

> *“I do feel a bit happier now, because I feel like the responsibilities, especially during the pandemic, and I felt like, although I had more responsibilities, I managed to fulfil them. So that sense of fulfilment and achievement that I felt, although I didn’t receive that much external support” (Young Carer 4, age between 22- 24, looks after parent)*

> *“I think it’s made me realise my own strength as a carer, during the pandemic. The fact that I could get through a really difficult time” (Young Carer 9, age between 18- 21, looks after parent)*

Whilst for others, the pandemic provided them with the opportunity to reflect on what was important to themselves and to implement strategies to achieve goals in relation to this, whether that be their own mental health and wellbeing:

> *“I got insecure and [started] working through self-harm issues because of being insecure, I completely changed the way I look and I completely stopped lying to myself about who I was and remembered who I want to be. And that has given me, now I have a lot more confidence. I still have issues with self-harm but I am very confident in the way I look and the way I feel about myself. I have security in knowing I can do it on my own if I need to” (Young Carer 6, age between 14- 17, looks after grandparent)*

or their education:

> *“So, my school chose not to give any materials to work on out, so in that six months, I just created a plan, a timetable, where I would work every single day in revising. It was a bit more independent on myself” (Young Carer 8, age between 18- 21, looks after multiple family members)*

## Discussion

### Main findings

We aimed to understand the impact of the pandemic and associated restrictions on mental health, wellbeing and access to support for Young Carers living in the UK, with a view of making suggestions to future service provision, as well as how Young Carers could be better supported in future health emergencies.

Whilst the pandemic had negative consequences for many young people (McKinlay et al., 2022), Young Carers have been disproportionally affected, with research highlighting increased mental health difficulties and lowered wellbeing when compared to peers with non-caring responsibilities (Landi et al., 2022; Nakanishi et al., 2022). Previous literature, as well as findings from this study, can help illuminate reasons why. In keeping with previous findings, for many Young Carers, having a routine was important as this helped them manage their daily caring responsibilities (Blake-Holmes, 2020). Similarly, school was seen as an important part of this routine as it provided a form of self-care and respite (Blake-Holmes, 2020). However, a novel finding from this study was that participants also outlined how school provided a place to access emotional support via school staff and links into other agencies. As schools went into lockdown and staff became overwhelmed with tasks such as adapting lessons for online teaching, this meant that Young Carers lost, or had limited access to this emotional support pathway. This, combined with losing other sources of support may have contributed to the increases in mental health difficulties and lowered wellbeing observed in other studies (Landi et al., 2022; Nakanishi et al., 2022). To support Young Carers in future, schools should actively enquire with students each year to understand which pupils may be classified as Young Carers. However, as some individuals do not know they fall under this classification (Darling et al., 2019), this should be handled in a sensitive way by form tutors or pastoral staff. Once identified, policies and procedures for protected time with Young Carers should be implemented to secure both their emotional wellbeing and support their learning, both in and out of health emergencies. Given the limited time within the school day and their caring responsibilities, possible considerations may include smaller class sizes, greater 1-1 support during the school day by teachers and pastoral staff or extending the school terms to include specific summer school programmes for Young Carers.

In addition to schools, many Young Carers reported peer support and friendship groups as important sources of emotional support as they allowed time for respite and to engage in similar activities to their peers. Practically, the closure of schools and pandemic related restrictions such as lockdowns meant that Young Carers were only able to see their friends digitally, which for some, resulted in the fragmenting of friendship groups, whose size did not easily adapt to these digital interactions, or disengagement from friendship groups due to zoom fatigue. For others, more caring responsibilities meant they had less time to engage with friends and peers. Results from a cohort study found that during the pandemic, increased distress and lower mental wellbeing were identified in Young Carers and driven by aspects such as low social support and feelings of isolation (Nakanishi et al., 2022).

Given the importance placed on friendship groups for Young Carers, our findings help shed further light on specific reasons for the breakdown of these social support structures. To protect Young Carers during future health emergencies, the Government may wish to consider creating support bubbles specifically involving groups of Young Carers, particularly as those we interviewed followed the rules because they did not want to put those they cared for at risk, which may class them as low risk of spreading the virus within those support bubbles.

An important finding identified from participant interviews in this study was the additional and enhanced forms of support provided by other agencies, predominantly local authorities and third sector organisations. Participants reported that these organisations not only provided emotional and wellbeing support but acted as a conduit to address their basic physiological and safety needs, facilitating access to resources such as funds and food parcels. This meant that for some Young Carers, they were able to concentrate on other priorities, such as additional caring responsibilities, or their education. However, whilst these forms of support were welcomed, emotional or peer group support sessions, which took place over videocall, were seen by many as suboptimal and cognitively taxing, particularly if their education was provided via this same medium. Similar to the previous paragraph, one solution to support the emotional wellbeing of Young Carers may be to allow local authorities or third sector organisations to facilitate or host in-person support bubbles specifically for Young Carers. However, given the importance of local authorities and third sector organisations ability to step in to support this group when other services were overwhelmed, the Government should consider ring fencing additional funds specifically for supporting the emotional wellbeing of Young Carers, to be made available not only during future health emergencies, but also more generally, given the lack of educational and mental health parity when compared to their peers without caring responsibilities (Becker & Sempik, 2019).

A further novel finding is the perceived difficulty in getting the individual(s) the Young Carer looked after to follow the rules, often due to cognitive or mental health difficulties. This caused distress to the Young Carers as it increased the possibility of the individuals being looked after, of who many were classified as vulnerable, catching the virus, which could be fatal. In future health emergencies, the Government, as well as local and regional public health experts should draw on the support of clinical and health psychologists to provide targeted messaging to Young Carers and their families, as well as practical strategies to support adherence to guidelines. In this instance, these targeted messages and support strategies may wish to draw on CBT-informed, and easy-to-understand communications for those with cognitive or mental health difficulties. Moreover, Psychologists may also want to consider implementing group sessions with Young Carers to help reinforce the messages they are trying to promote, as Young Carers will have a large responsibility in managing their looked after persons’ care. Similar targeted messaging drawing on behaviour change principles may also wish to be employed for immediate family members of Young Carers, however, a further understanding on commonalities regarding non-adherence to rules and guidelines with these individuals is needed.

Our sample was ethnically diverse, with nearly three quarters (71%) being from minority ethnic backgrounds. Whilst this proportion is higher than the national average (Neale, 2023) it provides important insights into underserved groups who are Young Carers and has implications on how these individuals can be supported. Recent reports have identified that Young Carers from Asian or mixed backgrounds are more likely to be unhappy across most aspects of life when compared to white young carers (Childrens Commissioner, 2022), as well as more being more likely to experience poor mental health (The Childrens Society, 2018). Given the importance of some specialist third sector organisations in supporting and building strong relationships with Young Carers, consideration should be given to developing pathways for Young Carers into services, with special attention being paid to Young Carers with intersectional characteristics, such as those who are from minority backgrounds.

This is particularly important as currently pathways into mental health services for minority ethnic young people are more likely to be from youth justice or social care settings and therefore compulsory in nature (Edbrooke-Childs & Patalay, 2019) which may cause stigma, remove choice and empowerment and lead to worse health outcomes (Ruphrect-Smith et al., 2023).

### Strengths and limitations

This study builds on initial research conducted with Young Carers at the start of the pandemic by allowing participants to reflect on later stages as restrictions eased and guidelines changed. Our research contributes an in-depth picture of the impact of the pandemic for Young Carers exploring a range of issues with participants and provides recommendations that can be carried forward to protect this group in future health emergencies. Our sample was ethnically diverse and represented a range of caring responsibilities and caring relationships. However, whilst a range of viewpoints were sought, this study relied on a convenience sample and thus viewpoints from certain demographics may be missing or under-represented. Moreover, we used remote interviewing techniques which may have excluded some groups with limited online access.

### Future research

Future research should focus on working with Young Carers, their families and organisations involved in their co-ordination and care to explore how best to put these recommendations into practice. Moreover, given that these recommendations were provided by academics as a result of interviews, future research should also explore directly with Young Carers other strategies which may be helpful in supporting Young Carers in future health emergencies. To translate findings into practice, research findings should incorporate implementation science or behaviour change frameworks to understand how to adapt and enhance access to existing programmes of support, and co-produce outputs for maximum reach and effectiveness. Future research should also consider perspectives from family members looked after by Young Carers, understand both their experiences during health emergencies, as well as their views on how to support both themselves and Young Carers in future.

## Conclusions

We suggest a range of interventions that could be implemented across different systems to support Young Carers in future health emergencies and make sure this group is not overlooked in future. From a Governmental perspective, ministers should consider guidelines and provisions to allow for support bubbles involving groups of Young Carers, so that they are able to provide emotional support to one-another, as well as ring fence funding specifically for this cohort, which should be used by local authorities and third sector organisations to provide physiological, social and emotional support. Third sector organisations and local authorities should develop policies and guidelines based on their learning to further streamline support for Young Carers in future healthcare emergencies but also in a post-pandemic world. Lastly, those responsible for public health, should consider working closely with psychologists and local authorities to create targeted messaging to reinforce health promoting behaviours for those with cognitively impairments or for wider family support networks of populations vulnerable to negative health impacts of the virus, and psychologists may wish to provide training and support to Young Carers to help reinforce such messages when at home with family members.

## Data Availability

The datasets generated and/or analysed during the current study are not publicly available, nor are they available upon request, because the dataset consist of interview transcripts that might compromise participant privacy and confidentiality due to the sensitive nature of topics discussed.

## List of abbreviations

GCSE: General Certificate of Secondary Education
GP: General Practitioner
OCD: Obsessive Compulsive Disorder
UCL: University College London
UK: United Kingdom

## Declarations

### Ethics approval and consent to participate

The UCL Ethics Committee reviewed and approved this study (Project IDs: 14895/005 and 6357/002). We confirm that all methods were carried out in accordance with relevant guidelines and regulations under ethics approval, including that all participants provided their informed assent/consent (and where appropriate parent/guardian consent) to participate in this research.

### Consent for publication

Not applicable

### Competing interests

None declared

### Funding

The Covid-19 Social Study was funded by the Nuffield Foundation [WEL/ FR-000022583], but the views expressed are those of the authors and not necessarily the Foundation. The study was also supported by the MARCH Mental Health Network funded by the Cross-Disciplinary Mental Health Network Plus initiative supported by UK Research and Innovation [ES/S002588/1], and by the Wellcome Trust [221400/Z/20/Z]. DF was funded by the Wellcome Trust [205407/Z/16/Z].

### Authors’ contributions

DF and AB conceived the study design. AB carried out data collection. DH led on data analysis with support from AB. DH wrote the first draft of the manuscript. All authors have read, provided revisions, and approved the final version of this manuscript.

## Acknowledgements

The authors would like to thank Dr Alison McKinlay for her insights into theme development and Young Carer organisations for their assistance with recruitment of participants for this study, as well as those individuals who gave up their time to take part and contribute to the study.

## Notes

### Competing Interest Statement

The authors have declared no competing interest.

### Author Declarations

All procedures involving human participants were approved by the University College London Ethics Committee (project IDs: 14895/005 and 6357/002).

## References

Becker, S., & Sempik, J. (2019). Young Adult Carers: The Impact of Caring on Health and Education. Children & Society, 33(4), 377–386. 10.1111/chso.12310

Blake-Holmes, K. (2020). Understanding the needs of Young Carers in the context of the Covid 19 Global Pandemic.

Blake-Holmes, K., & McGowan, A. (2022). ‘It’s making his bad days into my bad days’: The impact of coronavirus social distancing measures on young carers and young adult carers in the United Kingdom. Child and Family Social Work, 27(1), 22–29. 10.1111/cfs.12877

Braun, V., Clarke, V., & Hayfield, N. (2022). ‘A starting point for your journey, not a map’: Nikki Hayfield in conversation with Virginia Braun and Victoria Clarke about thematic analysis. Qualitative Research in Psychology, 19(2), 424–445. 10.1080/14780887.2019.1670765

Carers Trust. (2016). Invisible and in distress: Prioritising the mental health of England’s young carers. .

Carers Trust. (2020). About young adult carers. .

Chadi, N., & Stamatopoulos, V. (2017). Caring for young carers in Canada. Canadian Medical Association Journal, 189(28), E925–E926. 10.1503/cmaj.170145

Cheesbrough, S., Harding, C., Webster, H., Taylor, L., & & Aldridge, J. (2017). The lives of young carers in England. Omnibus survey report.

Children’s Commissioner. (2016). Young carers: The support provided to young carers in England. .

Childrens Commissioner. (2022). Young carers findings from The Big Ask. https://assets.childrenscommissioner.gov.uk/wpuploads/2022/03/cco-young-carers-findings-from-the-big-ask-march-22.pdf

Darling, P., Jackson, N., & Manning, C. (2019). Young carers: unknown and underserved. British Journal of General Practice, 69(688), 532–533. 10.3399/bjgp19X706121

Edbrooke-Childs, J., & Patalay, P. (2019). Ethnic Differences in Referral Routes to Youth Mental Health Services. Journal of the American Academy of Child & Adolescent Psychiatry, 58(3), 368–375.e1. 10.1016/j.jaac.2018.07.906

Fancourt, D., Steptoe, A., & Bu, F. (2021). Trajectories of anxiety and depressive symptoms during enforced isolation due to COVID-19 in England: a longitudinal observational study. The Lancet Psychiatry, 8(2), 141–149. 10.1016/S2215-0366(20)30482-X

Ganson, K. T., Tsai, A. C., Weiser, S. D., Benabou, S. E., & Nagata, J. M. (2021). Job Insecurity and Symptoms of Anxiety and Depression Among U.S. Young Adults During COVID-19. Journal of Adolescent Health, 68(1), 53–56. 10.1016/j.jadohealth.2020.10.008

Hovstadius, B., Ericson, L., & Magnusson, L. (2015). Barn Som Anhöriga: Ekonomisk Studie av Samhällets Långsiktiga Kostnader (Children as Next of Kin—An Economic Study of Society’s Long Term Costs).

Istat. (2023, January 4). Condizioni di Salute e Ricorso ai Servizi Sanitari in Italia e nell’Unione Europea—Indagine EHIS 2015. . Available Online: https://www.Istat.It/It/Archivio/204655.

James, E. (2017). Still Hidden, Still Ignored Who cares for young carers? https://www.barnardos.org.uk/sites/default/files/uploads/still-hidden-still-ignored.pdf

Joseph, S., Sempik, J., Leu, A., & Becker, S. (2020). Young Carers Research, Practice and Policy: An Overview and Critical Perspective on Possible Future Directions. Adolescent Research Review, 5(1), 77–89. 10.1007/s40894-019-00119-9

Landi, G., Pakenham, K. I., Grandi, S., & Tossani, E. (2022). Young Adult Carers during the Pandemic: The Effects of Parental Illness and Other Ill Family Members on COVID-19- Related and General Mental Health Outcomes. International Journal of Environmental Research and Public Health, 19(6), 3391. 10.3390/ijerph19063391

Lee, J. (2020). Mental health effects of school closures during COVID-19. The Lancet Child & Adolescent Health, 4(6), 421. 10.1016/S2352-4642(20)30109-7

Leu, A., & Becker, S. (2019). Young Carers. In Childhood Studies. Oxford University Press. 10.1093/obo/9780199791231-0120

Leu, A., Berger, F. M. P., Heino, M., Nap, H. H., Untas, A., Boccaletti, L., Lewis, F., Phelps, D., Santini, S., D’Amen, B., Socci, M., Hlebec, V., Rakar, T., Magnusson, L., Hanson, E., & Becker, S. (2022). The 2021 cross-national and comparative classification of in-country awareness and policy responses to ‘young carers.’ Journal of Youth Studies, 1–18. 10.1080/13676261.2022.2027899

Leu, A., Frech, M., Wepf, H., Sempik, J., Joseph, S., Helbling, L., Moser, U., Becker, S., & Jung, C. (2019). Counting Young Carers in Switzerland - A Study of Prevalence. Children & Society, 33(1), 53–67. 10.1111/chso.12296

Levine, C., Hunt, G. G., Halper, D., Hart, A. Y., Lautz, J., & Gould, D. A. (2005). Young Adult Caregivers: A First Look at an Unstudied Population. American Journal of Public Health, 95(11), 2071–2075. 10.2105/AJPH.2005.067702

Mansfield, R., Santos, J., Deighton, J., Hayes, D., Velikonja, T., Boehnke, J. R., & Patalay, P. (2022). The impact of the COVID-19 pandemic on adolescent mental health: a natural experiment. Royal Society Open Science, 9(4). 10.1098/rsos.211114

Martin, S. (2021). Young Carers and COVID-19: The Invisible Front-Line Workers. The University of Guelph.

McKinlay, A. R., May, T., Dawes, J., Fancourt, D., & Burton, A. (2022). ‘You’re just there, alone in your room with your thoughts’: a qualitative study about the psychosocial impact of the COVID-19 pandemic among young people living in the UK. BMJ Open, 12(2), e053676. 10.1136/bmjopen-2021-053676

Nakanishi, M., Richards, M., Stanyon, D., Yamasaki, S., Endo, K., Sakai, M., Yoshii, H., & Nishida, A. (2022). Adolescent Carers’ Psychological Symptoms and Mental Well-being During the COVID-19 Pandemic: Longitudinal Study Using Data From the UK Millennium Cohort Study. Journal of Adolescent Health, 70(6), 877–884. 10.1016/j.jadohealth.2022.01.228

Neale, B. (2023). Young carers and young adult carers from underrepresented groups: What do we know? Ethnic minority and LGB+ young carers and young adult carers. Young Carers. xhttps://carers.org/downloads/young-carers-from-under-represented-backgrounds---slides.pdf

QSR International Pty Ltd. (2018). NVivo (Version 12).

Ruphrect-Smith, H., Davies, S., Jacob, J., & Edbrooke-Childs, J. (2023). Ethnic differences in treatment outcome for children and young people accessing mental health support. European Child & Adolescent Psychiatry. 10.1007/s00787-023-02233-5

Sieh, D. S., Meijer, A. M., Oort, F. J., Visser-Meily, J. M. A., & Van der Leij, D. A. V. (2010). Problem Behavior in Children of Chronically Ill Parents: A Meta-Analysis. Clinical Child and Family Psychology Review, 13(4), 384–397. 10.1007/s10567-010-0074-z

Stamatopoulos, V. (2015). One million and counting: the hidden army of young carers in Canada. Journal of Youth Studies, 18(6), 809–822. 10.1080/13676261.2014.992329

Stamatopoulos, V. (2018). The young carer penalty: Exploring the costs of caregiving among a sample of Canadian youth. Child & Youth Services, 39(2–3), 180–205. 10.1080/0145935X.2018.1491303

Steinberg, L., & Morris, A. S. (2001). Adolescent Development. Annual Review of Psychology, 52(1), 83–110. 10.1146/annurev.psych.52.1.83

The Childrens Society. (2018). Young Carers of Black and Minority Ethnic families.

van Tienen, I., de Roos, S., & de Boer, A. (2020). Spijbelen onder scholieren: de rol van een zorgsituatie thuis. TSG - Tijdschrift Voor Gezondheidswetenschappen, 98(1), 9–17. 10.1007/s12508-020-00253-z

Wylie-Curia, N. (2021). “Now We Spend Every Waking Hour Together”: Experiences of Young Carers in the COVID-19 Pandemic. The University of Guelph.

Young Minds. (2021). Coronavirus: Impact on young people with mental health needs. [Internet], 2021. Available:

